# Clinical Characteristics of Hospitalized Patients with SARS-CoV-2 and Hepatitis B virus Co-infection

**DOI:** 10.1101/2020.03.23.20040733

**Authors:** Xiaoping Chen, Qunqun Jiang, Zhiyong Ma, Jiaxin Ling, Wenjia Hu, Qian Cao, Pingzheng Mo, Rongrong Yang, Shicheng Gao, Xien Gui, Yong Xiong, Jinlin Li, Yongxi Zhang

## Abstract

**Background & Aims:** The coronavirus disease 2019 (COIVD-19) caused by SARS-CoV-2 has been characterized as a pandemic, which causes a serious public health challenge in the world. A very large group of patients infected by HBV has been reported worldwide, especially in China. In order to answer whether specific treatment strategy on the patients coinfected with HBV and SARS-CoV-2, it requires profound understanding of the clinical characteristics on those patients. However, the impacts of SARS-CoV-2 infection on HBV patients remain largely unknown.

**Approach & Results:** In this retrospective investigation, we included 123 COVID-19 patients admitted to Zhongnan Hospital of Wuhan University, Wuhan, China, from January 5 to March 7, 2020. All enrolled patients are the laboratory confirmed COVID-19 pneumonia cases according to the criteria reported previously. A total of 123 patients were analyzed for their Clinical records, laboratory results including the diagnosis of HBV infection and liver function. Among 123 confirmed COVID-19 patients, the mean age was 51 years old and 59.3% were females (73/123). Fifteen were previously HBV infected patients, 66.7% of them were males (10/15), patients with HBV infection appeared to have a higher incidence of liver cirrhosis and an increased level of total bilirubin. Seven (46.7%) patients with HBV infection were defined as severe cases, while the severity rate was 24.1% for the patients without HBV infection (26/108). The mortality of patients with HBV infection was 13.3% (2/15) compared to 2.8% (3/108) for the patients without HBV infection.

**Conclusions:** SARS-CoV-2 infection may cause Live function damage in COVID-19 cases and the patients with HBV infection are likely to have more severe disease outcome.

---

In early December 2019, there was an outbreak of novel coronavirus-associated pneumonia in Wuhan, China. The virus was spreading rapidly to other cities of China and accumulating cases had been reported in coming days(1). According to the announcement of the World Health Organization (WHO), the disease has been officially named as Coronavirus Disease-2019 (COVID-19) (2). The etiology of the disease was identified to be a novel β-coronavirus, named as severe acute respiratory syndrome coronavirus 2 (SARS-CoV-2) based on the phylogenetic relationship with SARS-CoV. On March 11, 2020, WHO declared the outbreak of SARS-CoV-2 as a pandemic. So far, more than 290,000 people in over 180 countries or territories have reported COVID-19 cases, and more than 12,000 people have died according to data from WHO (https://www.who.int/emergencies/diseases/novel-coronavirus-2019). Around 25% COVID-19 cases were reported in Wuhan, China.

In addition to the recent emerged SARS-CoV-2, Hepatitis B virus (HBV) is one of the viruses which causes a global infection and threat public health. In worldwide, the prevalence of HBsAg is about 3.9%. As high as 290 million patients are suffering from chronic HBV infection and about 650,000 patients die from HBV infection due to liver failure, liver cirrhosis and hepatocellular carcinoma (HCC) each year(3, 4). According to a nationwide epidemiological survey of population whose ages range from 1 to 59y in China, 2016, the prevalence of HBsAg was 7.2%. Around 93 million patients were positive for HBV infection and 20 million patients were diagnosed as chronic hepatitis B infection(5, 6).

Previous studies have shown that SARS-CoV-2 has a capacity to infect multiply organs including upper respiratory tract, lung, kidney probably due to the expression of SRAS-CoV-2 receptor, Angiotensin-converting enzyme 2 (ACE2), on these tissues(7). A recent research has demonstrated that SARS-CoV-2 infection was associated with live function damage in COVID-19 patients(8). Taking consideration of large group of people with HBV infection, the risk of SARS-CoV-2 infection on patients with HBV infection requires a further assessment in order to design the specific treatment strategy. However, the impacts of SARS-CoV-2 infection on HBV patients are still not clear. For example, we do not yet know whether the SARS-CoV-2 infection is more severe in HBV patients and we also do not have much knowledge about the impact of SARS-CoV-2 on the course of HBV infection. In this retrospective study, we discovered that the liver impairment is a common feature in COVID-19 patients and as high as 46.7% patients with HBV infection develop to severe situation during the course of SARS-CoV-2 infection. This suggests that patients with HBV infection might be vulnerable group to SARS-CoV-2 infection.

## Methods

## Study design

From January 5 to February 7, 2020, 123 COVID-19 patients were enrolled in the study. Informed consents were obtained from all patients upon admission to the Department of Infectious Diseases, Zhongnan Hospital of Wuhan University, Wuhan, China. The clinical outcomes (ie, discharges, mortality, Hospital stays) were monitored up to March 7, 2020, the final date of follow-up.

### Data collection

The information of enrolled patients including the demographic information, clinical manifestations, laboratory data including blood routine examination, liver function, Hepatitis B virus serological markers (HBsAg, anti-HBsAg, HBeAg, anti-HBeAg, anti-HBcAg, HBV-DNA), and outcome of disease, were collected and reviewed by two researchers to avoid subjective biases.

The diagnosis of COVID-19 was based on real-time RT-PCR. Throat swab samples were collected for extracting SARS-CoV-2 RNA from patients suspected of having SARS-CoV-2 infection as described anywhere(9). The diagnostic criteria of SARS-CoV-2 real-time RT-PCR were based on the recommendation by the National Institute for Viral Disease Control and Prevention, China (http://ivdc.chinacdc.cn/gjhz/jldt/202002/P020200209712430623296.pdf).

Severe patients were defined according to the Guideline of the treatment of COVID-19 (Version 6, 2020 Feb 18, http://www.nhc.gov.cn/yzygj/s7653p/2020028334a8326dd94d329df351d7da8aefc2.shtml). Briefly, we categorize the patient as severe case if the symptoms of dyspnea show. The signs of dyspnea include any of the following features: shortness of breath, respiration rate ≥ 30bpm, blood oxygen saturation ≤ 93% (at rest), PaO2 / FiO2 ≤ 300 mmHg, or pulmonary inflammation that progresses dramatically within 24 to 48 hours> 50%.

### Statistical analysis

The statistical analyses in this study was performed by the SPSS 17.0 software package. We utilized *X*^*2*^ tests or Fisher’s exact tests for categorical variables. For normal distribution, *t-test* was applied to analyze the data, expressed as mean ± standard deviations. Regarding the non-normal distribution data, we used the Mann-Whitney U to do the test and the results were shown as of median (25%−75% interquartile range, IQR), A *p* value of < 0.05 was considered statistically significant.

### The principle of medical ethics

This study was approved by the ethics board in Zhongnan Hospital of Wuhan University, Wuhan, China (No.2020011).

## Results

### Baseline characteristics of COVID-19 patients with or without HBV infection

A total of 123 patients with COVID-19 were enrolled in this study, including 50 males and 73 females. Around 12.2% (15/123) of patients are also suffering from HBV infection. Males take up 66.7% (10/15) of patients coinfected by HBV and SARS-CoV-2 and seems to have a higher coinfection rate compared to females (p=0.0469, Table 1). The median age of total enrolled patients was 51.0 years (IQR, 35.0-66.0; range, 20-96 years). The most common symptoms at the onset of illness were: fever (37.4−39.1°C, 69.1%), fatigue (54.5%), cough (50.4%), myalgia (32.5%), and less common: dyspnea (21.1%), Headache (16.3%) and diarrhea (17.1%). Among the 123 patients, thirty-five (28.5%) cases had underlying at least one comorbidity such as hypertension, cardiovascular disease, diabetes, malignancy, COPD and liver cirrhosis. Patients with HBV infection had a higher rate of liver cirrhosis (p=0.0390, Table 1). Seven of 15 patients (46.7%) with HBV infection develop to the severe situation, while the percentage of severe cases is much less (24.1%) in the COVID-19 patients without HBV infection.

**Table 1.**
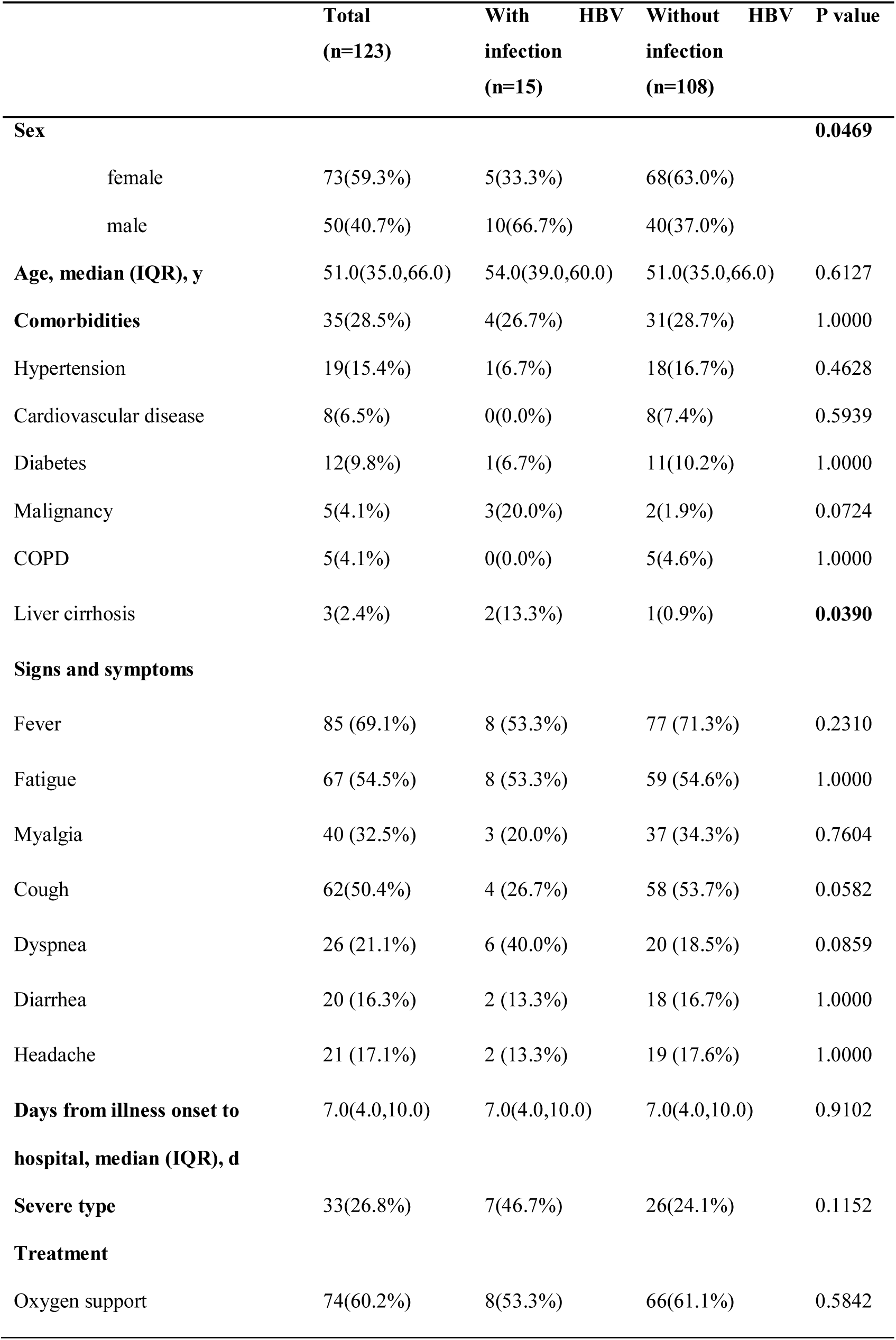
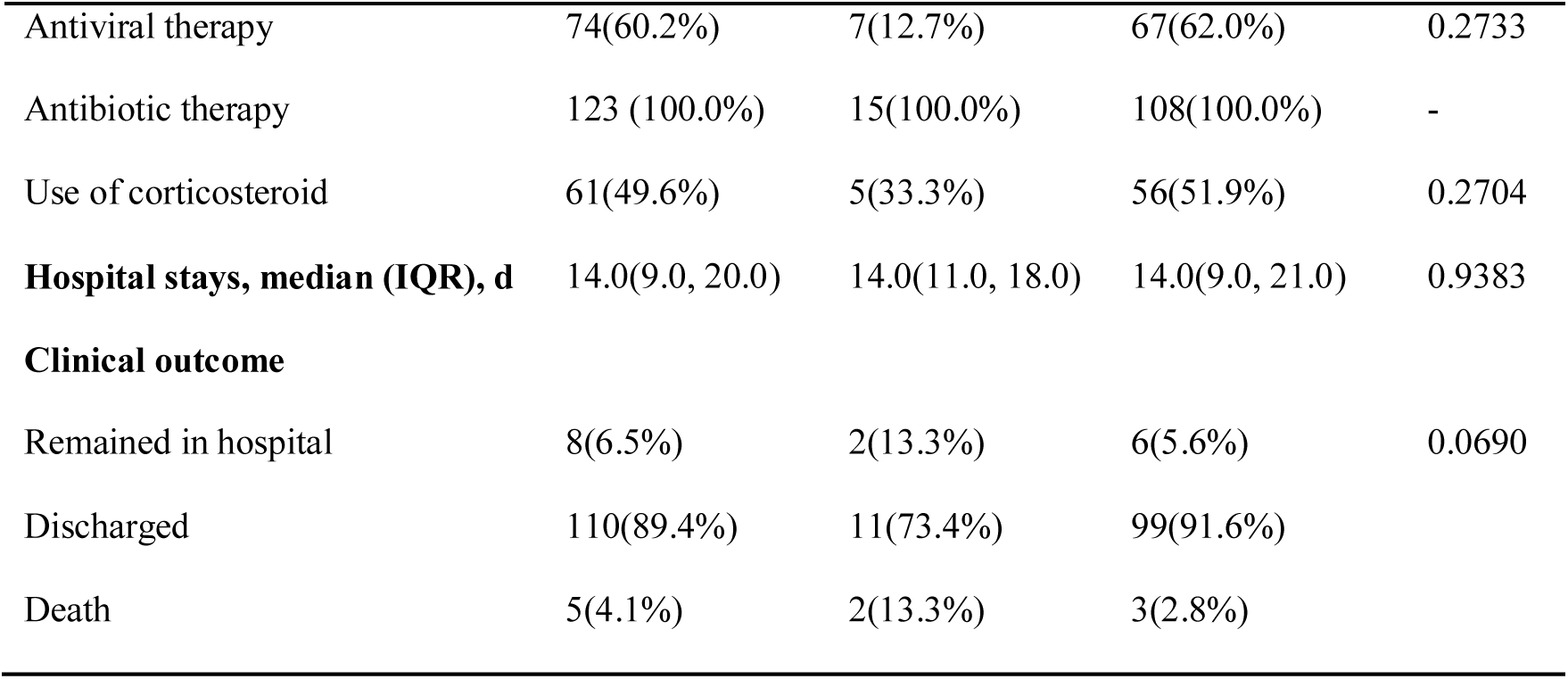
Demographics, baseline characteristics, treatment and clinical outcomes of 123 COVID-19 patients with or without HBV infection

The treatment was mainly the supportive care (Table 1). Seventy-four patients were given antiviral (arbidol, orally, 200 mg, three times per day), and 74 with oxygen support. Antibiotic therapy, both orally and intravenous, were given as described in Table 1. Sixty-one patients received corticosteroids to suppress an excessive inflammatory activation. There is no significant difference of treatment between patients with or without HBV infection.

### laboratory Findings of COVID-19 patients with or without HBV infection at baseline

The biochemical tests included measuring the level of alanine aminotransferase (ALT), aspartate aminotransferase (AST), total bilirubin (TBIL), gamma-glutamyltransferase (GGT), alkaline phosphatase (ALP), albumin as well as recording prothrombin time, activated partial thromboplastin time, international normalized ratio, d-dimer and creatinine. All of these biochemical features were found normal; however, the level of total bilirubin was higher in patients with HBV infection (p=0.0178, Table 2). The blood counts of the patients with or without HBV infection showed lymphopenia (< 1.3 ×10^9^/L).

**Table 2.**
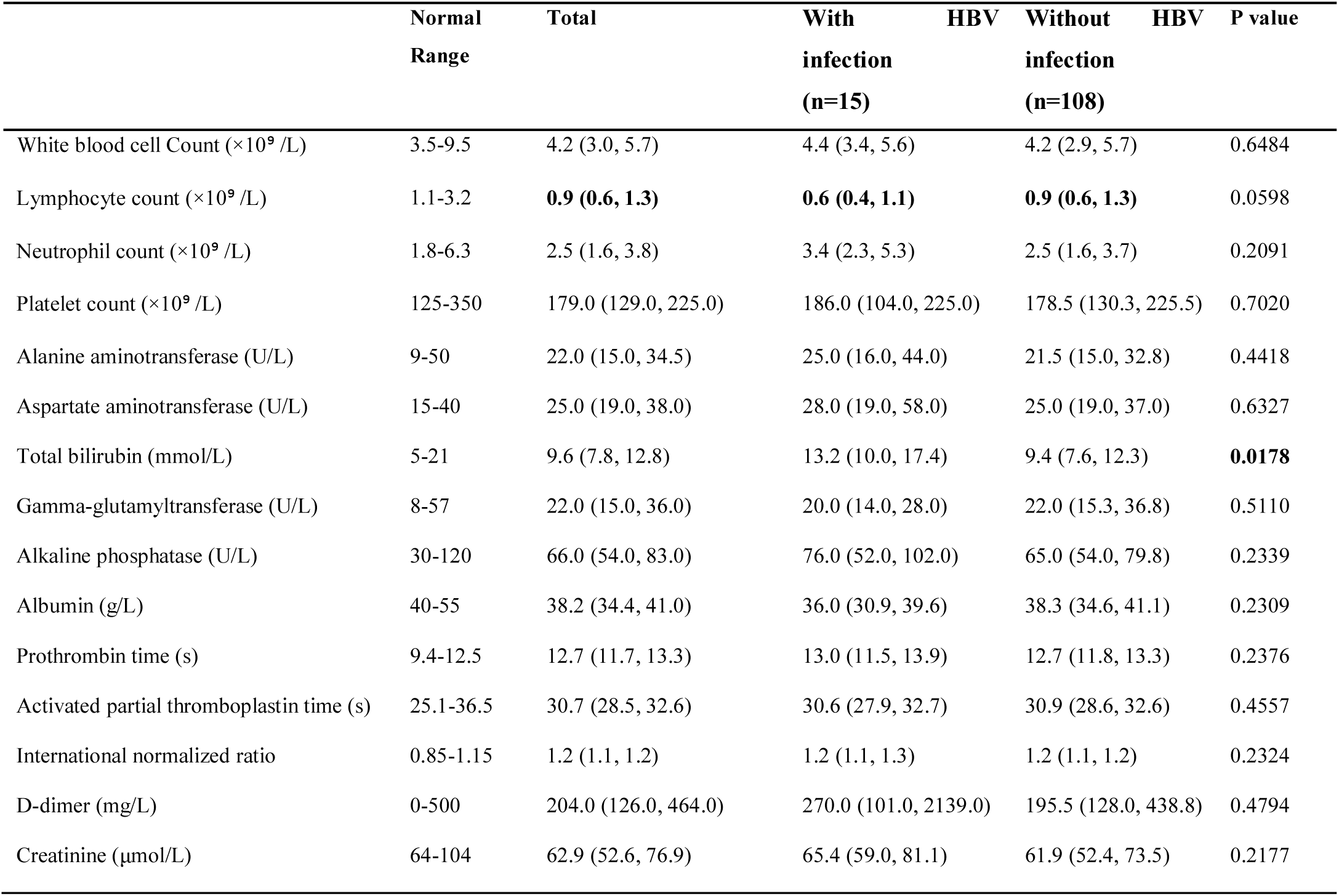
Laboratory results of 123 COVID-19 patients with or without HBV infection

### Hepatitis B serological markers of COVID-19 patients with HBV infection

Fifteen COVID-19 patients were examined to be HBsAg positive (5 females and 10 males). The data of anti-HBsAg, HBeAg, anti-HBeAg and anti-HBcAg were available for 11 patients with ten patients HBeAg negtive and one positive. The value of HBV-DNA was collected from 13 patients. The HBV-DNA level of 10 patients are more than 20 IU/ml (Table S1).

### Clinical outcome

We observed the clinical outcome of 123 COVID-19 patients within 31 days of treatment. Eleven patients (73.4%) with HBV infection and 99 patients (91.6%) without HBV infection were discharged from the hospital according to the guideline. Two patients (13.3%) with HBV infection and 6 patients (5.6%) without HBV infection were still hospitalized. Two patients (13.3%) with HBV infection and 3 patients (2.8%) without HBV infection were dead. Patients with HBV infection showed higher mortality rate compared to those COVID-19 patients without HBV infection (13.3%vs 2.8%, Table 2).

## Discussion

Resemble to the other two coronaviruses, SARS-CoV and MERS-CoV, SARS-CoV-2 can cause patients severe respiratory symptoms and even leads to death with average mortality rate of 3.4% (according to the data reported from WHO) though most cases of COVID-19 are acute and resolve fast. Liver damage has been identified in around 60% of patients suffering from SARS and viral RNA was detected by RT-PCR in liver tissue(10), which providing the evidence that SARS-CoV involved in liver injure. Liver impairment has been also reported in MERS patients(11). According to the clinical reports from different centers with large scale of COVID-19 cases, SARS-CoV-2 has been found to be associated with damage or dysfunction of liver tissue(9, 12-18) and about 14% - 53% COVID-19 cases showed liver function damage with abnormal level of alanine aminotransferase (ALT) and aspartate aminotransferase (AST). Our study is in line with previous observations. We found in COVID-19 cases without HBV infection that about 50.9% (55/108) patients have the dysfunction of liver symptoms by measuring the level of ALT, AST, TBIL, GGT, and ALP during the disease progress. In our enrolled cases, we also discovered that there is higher incidence of abnormal liver function (81.8%, 27/33) in severe COVID-19 patients than did in mild cases (43.3%, 39/90, data not shown), which agrees with the study that lower incidence of AST abnormality was found in the cases diagnosed by CT scan on the subclinical stage than in the COVID-19 patients who were confirmed after onset of symptom(15). Therefore, liver function could be considered as one factor to indicate the progress of COVID-19.

According to other study from 1099 cases, around 23.7% of confirmed COVID-19 patients have at least one comorbidity(13). Among these pre-existing chronic diseases, abnormal liver function is one of most common features in COVID-19 patients and severe patients are more likely to have HBV infection. In our research, about one out of five (7/33, 21.8%) COVID-19 severe patients were found to coinfect with HBV infection. It has been suggested that liver impairment in COVID-19 patients could be due to the virus direct attack or resulted by other causes such as drug toxicity and systemic inflammation(18). To detect the viral RNA and viral particles from liver biopsies of COVID-19 patients will be helpful to elucidate if virus infect liver tissue. Our results pointed out that as high as around 50% (7/15) of HBV patients were identified as severe COVID-19 cases. It is more likely that HBV patients will suffer from more severe situation during the disease progress when were encountered with SARS-CoV-2 infection. In our enrolled cases, two patients with SARS-CoV and HBV coinfection died on admission. One patient died from severe liver disease, haptic sclerosis. And the other died from intestinal hemorrhage, which seems to be associated the impairment of gastrointestinal tract. More coinfection cases analysis are required to further understand whether SARS-CoV-2 infection aggerates the progress of pre-existing disease and thereby cause death. There are different phases for HBV chronic infection including immunotolerant, viral suppression under long-term treatment with nucleotide analogues. In our current study, we collected the data of HBV on 15 coinfection patients at one time point, which were mainly used to identify HBV infection. More coinfection cases analysis is required to provide further evidences for evaluating the effects of SARS-CoV-2 infection on active HBV replication and live impairment at different time points for the HBV patients in different phases.

In conclusion, by respectively analyzing the patients with coinfection of SARS-CoV-2 and HBV, we found that the patients with pre-existing HBV infection will be much more vulnerable to SARS-CoV-2 infection. During the pandemic of SARS-CoV-2 infection, HBV patients should be given the specific protection.

## Data Availability

All data referred to in the manuscript are availability.

## Abbreviations

COVID-19: Coronavirus Disease-2019
HBV: Hepatitis B virus

---

**Table S1.** Hepatitis B serological markers of fifteen COVID-19 patients with HBV infection

| Demographics |  |  | Hepatitis B virus serological markers |  |  |  |  |  |
| --- | --- | --- | --- | --- | --- | --- | --- | --- |
| Patient | Age<br>(Year) | Sex<br>(Female/Male) | HBsAg<br>(IU/L,0-0.05) | Anti-HBsAg<br>(IU/L,0-10) | HBeAg<br>(s/co,0-1) | Anti-HBeAg<br>(s/co,>1) | Anti-HbcAg<br>(s/co,0-1) | HBV-DNA<br>(IU/L,<20) |
| 1 | 38 | Male | > 250.0 | NA* | NA | NA | NA | 100.0 |
| 2 | 54 | Male | 425.1 | NA | NA | NA | NA | NA |
| 3 | 74 | Male | 1.8 | 0.00 | 0.40 | 0.03 | 11.31 | < 20 |
| 4 | 36 | Female | 1294.0 | 0.07 | 0.95 | 1.11 | 10.72 | 211.0 |
| 5 | 48 | Male | > 250.0 | 0.10 | 0.47 | 0.01 | 12.02 | 235.0 |
| 6 | 60 | Male | 1.1 | 0.74 | 0.44 | 0.03 | 9.67 | <20 |
| 7 | 72 | Female | 558.7 | 0.29 | 0.40 | 0.01 | 11.58 | 40500.0 |
| 8 | 56 | Female | 148.9 | 0.23 | 0.37 | 0.01 | 10.74 | 40.6 |
| 9 | 57 | Male | 122.7 | NA | NA | NA | NA | NA |
| 10 | 39 | Male | > 250.0 | NA | NA | NA | NA | 657.0 |
| 11 | 50 | Female | 2971.0 | 0.00 | 0.38 | 0.02 | 10.39 | 2180.0 |
| 12 | 49 | Male | 143.9 | 0.77 | 0.40 | 0.02 | 10.85 | 89.0 |
| 13 | 59 | Male | 0.2 | 0.38 | 0.01 | 0.01 | 10.39 | <20 |
| 14 | 77 | Male | 5.6 | 5.14 | 0.42 | 0.07 | 10.90 | 166.0 |
| 15 | 28 | Female | > 250.0 | 0.00 | 2.60 | 1.32 | 6.44 | 1340.0 |

\*NA, not available.

## References

1. Li Q, Guan X, Wu P, Wang X, Zhou L, Tong Y, Ren R, et al. Early Transmission Dynamics in Wuhan, China, of Novel Coronavirus-Infected Pneumonia. N Engl J Med 2020.

2. Naming the coronavirus disease (COVID-2019) and the virus that causes it. In; 2020.

3. Ott JJ, Stevens GA, Groeger J, Wiersma ST. Global epidemiology of hepatitis B virus infection: new estimates of age-specific HBsAg seroprevalence and endemicity. Vaccine 2012;30:2212–2219.

4. Polaris Observatory C. Global prevalence, treatment, and prevention of hepatitis B virus infection in 2016: a modelling study. Lancet Gastroenterol Hepatol 2018;3:383–403.

5. In: Guidelines for the Prevention, Care and Treatment of Persons with Chronic Hepatitis B Infection. Geneva, 2015.

6. Chinese Society of Hepatology CMA, Chinese Society of Infectious Diseases CMA, Hou JL, lai W. [The guideline of prevention and treatment for chronic hepatitis B: a 2015 update]. Zhonghua Gan Zang Bing Za Zhi 2015;23:888–905.

7. Zou X, Chen K, Zou J, Han P, Hao J, Han Z. Single-cell RNA-seq data analysis on the receptor ACE2 expression reveals the potential risk of different human organs vulnerable to 2019-nCoV infection. Front Med 2020.

8. Zhenyu Fan LC, Jun Li, Cheng Tian, Yajun Zhang, Shaoping Huang, Zhanju Liu, Jilin Cheng. Clinical Features of COVID-19 Related Liver Damage. medRxiv 2020.02.26.20026971; 2020.

9. Wang D, Hu B, Hu C, Zhu F, Liu X, Zhang J, Wang B, et al. Clinical Characteristics of 138 Hospitalized Patients With 2019 Novel Coronavirus-Infected Pneumonia in Wuhan, China. JAMA 2020.

10. Chau TN, Lee KC, Yao H, Tsang TY, Chow TC, Yeung YC, Choi KW, et al. SARS-associated viral hepatitis caused by a novel coronavirus: report of three cases. Hepatology 2004;39:302–310.

11. Alsaad KO, Hajeer AH, Al Balwi M, Al Moaiqel M, Al Oudah N, Al Ajlan A, AlJohani S, et al. Histopathology of Middle East respiratory syndrome coronovirus (MERS-CoV) infection - clinicopathological and ultrastructural study. Histopathology 2018;72:516–524.

12. Chen N, Zhou M, Dong X, Qu J, Gong F, Han Y, Qiu Y, et al. Epidemiological and clinical characteristics of 99 cases of 2019 novel coronavirus pneumonia in Wuhan, China: a descriptive study. Lancet 2020;395:507–513.

13. Guan WJ, Ni ZY, Hu Y, Liang WH, Ou CQ, He JX, Liu L, et al. Clinical Characteristics of Coronavirus Disease 2019 in China. N Engl J Med 2020.

14. Huang C, Wang Y, Li X, Ren L, Zhao J, Hu Y, Zhang L, et al. Clinical features of patients infected with 2019 novel coronavirus in Wuhan, China. Lancet 2020:395:497–506.

15. Shi H, Han X, Jiang N, Cao Y, Alwalid O, Gu J, Fan Y, et al. Radiological findings from 81 patients with COVID-19 pneumonia in Wuhan, China: a descriptive study. Lancet Infect Dis 2020.

16. Xu XW, Wu XX, Jiang XG, Xu KJ, Ying LJ, Ma CL, Li SB, et al. Clinical findings in a group of patients infected with the 2019 novel coronavirus (SARS-Cov-2) outside of Wuhan, China: retrospective case series. BMJ 2020:368:m606.

17. Yang X, Yu Y, Xu J, Shu H, Xia J, Liu H, Wu Y, et al. Clinical course and outcomes of critically ill patients with SARS-CoV-2 pneumonia in Wuhan, China: a single-centered, retrospective, observational study. Lancet Respir Med 2020.

18. Zhang C, Shi L, Wang FS. Liver injury in COVID-19: management and challenges. Lancet Gastroenterol Hepatol 2020.

